# Mild Traumatic Brain Injury Disrupts Functional Dynamic Attractors of Healthy Mental States

**DOI:** 10.1101/19007906

**Authors:** Victor M. Vergara, Harm J. van der Horn, Andrew R. Mayer, Flor A. Espinoza, Joukje van der Naalt, Vince D Calhoun

## Abstract

The human brain has the ability of changing its wiring configuration by increasing or decreasing functional connectivity strength between specific areas. Variable but recurring configuration patterns in dynamic functional connectivity have been observed during resting fMRI experiments, patterns which are defined as dynamic brain states. The question arises whether in a regular healthy brain these states evolve in a random fashion or in a specific sequential order. The current work reveals both the specific state sequence in healthy brains, as well as the set of disruptions in this sequence produced by traumatic brain injury. The healthy sequence consists of oscillatory dynamic connectivity patterns that orbit an attractor state in a high dimensional space. Using discovery (96 subjects) and replication (74 subjects) cohorts, this study demonstrated that mild traumatic brain injury results in immediate orbital disruptions that recover over time. Brain dynamics enter a status of disrupted orbits right after injury, with partial recovery at 4 weeks, and full recovery at 3 months post-injury. In summary, our results describe an aspect of neuronal dysfunction in mild traumatic brain injury that is fully based on brain state dynamics, and different from traditional brain connectivity strength measures.

## 1. Introduction

Symptoms resulting from mild traumatic brain injury (mTBI) produce deleterious effects on cognitive and social functioning, and these deficits may last a lifetime for a minority of patients. Commonly reported symptoms include dizziness, vertigo, irritability, chronic headaches, difficulty concentrating, depression, and impulsiveness (DeKosky *et al*., 2010). The fact that these symptoms are notoriously difficult to objectify using current clinical measures provides a strong rationale for the ongoing research that tries to uncover the different aspects of this disease (Levin and Diaz-Arrastia, 2015). Previous studies suggested that functional magnetic resonance imaging (fMRI) is a promising technique to provide biomarker features (Vergara *et al*., 2017). Specifically, we showed that mTBI patients can be individually differentiated from healthy controls (HC) participants with high accuracy using dynamic functional network connectivity (dFNC), a measurement obtained from fMRI imaging (Vergara *et al*., 2018). However, previous analyses did not make full use of the temporal information in dFNC, limiting the method to extract features based on assessments of connectivity strength. Studies of patterns of functional connectivity transitions occurring in the brain are still an underexplored area. We predict that observed connectivity differences in mTBI compared to HC are rooted in patterns of dynamic transitions.

The varied accelerations of the head during a mTBI event affect the microstructure of axons, resulting in varying patterns of deficient anatomical connectivity (Huisman *et al*., 2004; Holli *et al*., 2010; Ling *et al*., 2012; Arenth *et al*., 2014). In accordance with white matter damage, abnormal patterns of functional connectivity have been found in several regions of the brain (Hillary *et al*., 2014; Sharp *et al*., 2014; Mayer *et al*., 2015a). Functional connectivity exists when there is temporal synchronicity of neuronal activation between two brain areas. Assessments of functional connectivity throughout the literature indicate a consistent map of functional connectivity abnormalities in mTBI. Resting state studies points towards the default mode network (DMN) as one of the main affected brain networks displaying a common pattern of weaker connectivity with other brain networks (Sours *et al*., 2013; Vakhtin *et al*., 2013; Sharp *et al*., 2014; Palacios *et al*., 2017). In contrast, increased functional connectivity within the DMN has been observed in some cases (Nathan *et al*., 2015; Vergara *et al*., 2017). Another consistent observation is increased functional connectivity that involves the cerebellum (Pagulayan *et al*., 2018), and especially between the cerebellum and the supplementary motor area (Nathan *et al*., 2015; Vergara *et al*., 2017; Vergara *et al*., 2018). However, effects in other networks may seem to differ. In the executive control network (ECN) increases (Borich *et al*., 2015) and decreases (Palacios *et al*., 2017) of functional connectivity has been reported. While reduced subcortical connectivity has been reported in mTBI (Vakhtin *et al*., 2013), thalamic connectivity has also been observed to increase (Tang *et al*., 2011). All of these observations have been made by considering that functional connectivity within a specific time lapse can be described by a single summary assessment. In such cases functional connectivity is assumed to have a quasi-static behavior with small variations that can be averaged (Allen *et al*., 2014). However, challenging this assumption might lead to an improvement of our understanding of brain mechanisms linked to TBI

Considering functional connectivity as a time evolving brain feature leads to several hypotheses that need to be described. As a preamble, we must view the brain as a dynamic system which is not always functionally connected in the same way in spite of a time invariant anatomical wiring. It is known that the human brain iterates among several patterns, also known as states, that can be identified using a dynamic functional connectivity method (Allen *et al*., 2014). Figure 1a shows an example of two different states and the two assumptions (Sharp and Smooth dynamic transitions) focus of this discussion. The simplest model describes the connectivity as unchanged through the duration of a state followed by a jump to the next state. While it is known that time variations of connectivity occur, no assessment of temporal nature is made within the state (Miller *et al*., 2016). We believe a more realistic assumption is a time-related variation of connectivity strength within the state, which is our first hypothesis. Figure 1a presents this alternative concept in a simplified manner. Second, we hypothesize the existence of oscillations of weak and strong connectivity within particular states as exemplified in Figure 1b. We propose to model these oscillations as a brain connectivity state that orbits around a given attractor connectivity pattern. The center of such an orbit is related to the state centroids as some examples have been previously identified in mTBI (Vergara *et al*., 2018). To represent the attractor and orbit within a state, we have chosen to display a double arrow from weak to strong and back. The third hypothesis is that attractor states are part of the normal brain behavior belonging to healthy controls as shown in Figure 1c. States not exhibiting oscillations may also be present, representing an intermediate transition between attractor states. Finally, we hypothesize that mTBI can disrupt the attractor states in such a way that no oscillation is observed. This abnormality is illustrated in Figure 1d. Different from previous assessments, these disruptions may not be directly related to connectivity strength and may only be observed when using the more dynamic attractor model.

**Figure 1.**
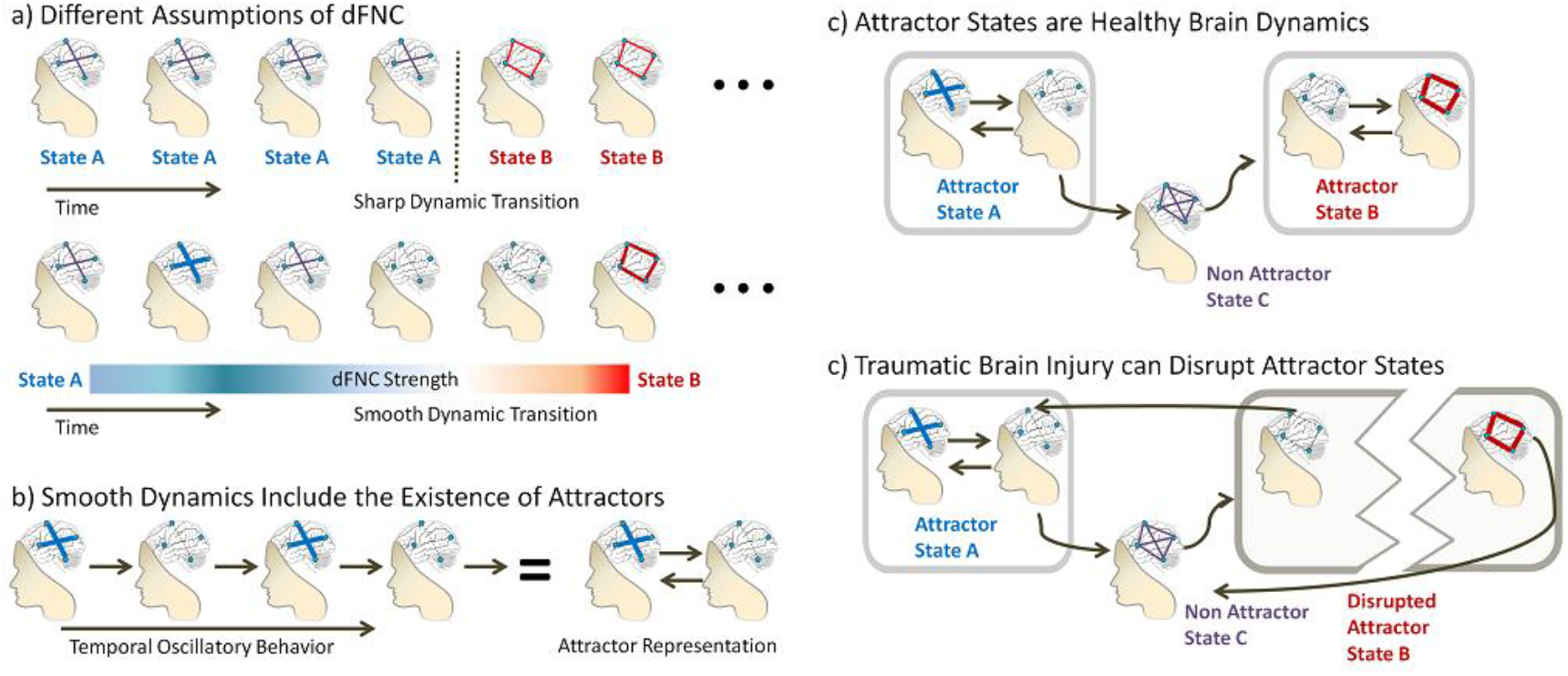
Comparison of a sharp transition assumption and a smooth transition assumption in dynamic functional network connectivity (dFNC). Part a) illustrates the differences between an assumption of time lapses with quasi-constant behavior and sharp dFNC state transitions versus a non-static connectivity change where the transition is smoother. Part b) shows that instead of transitioning, the state can start a dFNC oscillation without changing the state. The connectivity pattern is termed an attractor because it pulls the brain to remain in a specific dFNC state within a particular dynamicity. Part c) shows the hypothesis that attractors are part of a healthy brain behavior. A traumatic brain injury event can disrupt the attractor leading to abnormal brain dynamicity as illustrated in d).

This study examined the existence of attractor states in the healthy brain and attractor disruptions in subjects with mTBI. Two sample cohorts acquired by two different research projects were used in a discovery (96 subjects) and replication (123) scheme. A modified version of dynamic functional network connectivity (dFNC) (Allen *et al*., 2014) was applied that includes the first order derivatives of the time varying connectivity (Espinoza *et al*., 2018). Time derivatives impose an additional temporal constraint for the dFNC algorithm aiding to identify the different attractor sides illustrated in Figure 1.

## 2. Materials and methods

Two sample cohorts were used in this study. The discovery cohort was obtained in New Mexico and used for hypothesis testing. The replication cohort was obtained in the Netherlands and used to validate the results. A complete description of scanned subject and data preprocessing for both cohorts is provided in the supplementary documentation.

### 2.1. Discovery Cohort

We performed eyes-open resting state fMRI scans in 48 subjects with mTBI subjects and 48 age and sex matched healthy controls. Cohort’s mean age = 27.3 ± 9.0 years. The mTBI patients went through clinical (mean day post-injury = 13.9 ± 4.9 days) and brain imaging (mean day post-injury = 14.0 ± 5.3 days) evaluations within 21 days of injury. These 96 scans have been previously used in several studies, and extensive details of the clinical assessments can be found in the corresponding publications (Mayer *et al*., 2011; Mayer *et al*., 2015b; Vergara *et al*., 2017). Scans were used for dFNC analysis similar to a previous publication, where four dynamic states were identified and a total of 48 brain networks of interest (NOIs) were used (Vergara *et al*., 2018). The 48 NOIs were grouped in nine different functional connectivity domains including subcortical (SBC), cerebellum (CER), auditory (AUD), sensorimotor (SEN), visual (VIS), salience (SAL), default mode network (DMN), executive control network (ECN) and language (LAN). We provided spatial maps of these NOIs in the Supplementary Figure. We will focus on the time varying dynamic connectivity matrices of these NOIs. The dynamic analysis results in one connectivity matrix per time point. Current preprocessing differs from the previous publication (Vergara *et al*., 2018) as we used an artifact reduction connectivity assessment termed average sliding window correlation (ASWC) (Vergara *et al*., 2019) as well as temporal derivatives of ASWC (DASWC). In summary, there is one connectivity matrix (ASWC) and one connectivity derivative matrix (DASWC) for each time point.

The dFNC analysis identified lapses of time with a similar connectivity (ASWC) or derivative (DASWC) patterns using a k-means algorithm. In the context of the clustering output there are six clusters corresponding to six different derivative patterns. As displayed in Figure 2, there are only four connectivity patterns since cluster 1 and 2 exhibit the same whole-brain connectivity, despite of having different derivatives, where the DMN is negatively correlated with other cortical areas. This negative correlation is known to be the main characteristic of a resting state experiment (Greicius *et al*., 2003; Greicius *et al*., 2009; Shirer *et al*., 2012), but this is the first time that such pattern is observed split between two clusters. We say that whole-brain connectivity is in State A when the brain is in this connectivity pattern occurring during clusters 1 and 2. In a similar manner, State B is represented by the connectivity pattern of clusters 3 and 4 where in this case the SBC domain is negatively correlated with other cortical areas. Although it has not been the focus of resting state studies, this negative correlation between cortical and SBC domains is common occurrence in dFNC analysis (Allen *et al*., 2014). Matching of connectivity patterns of clusters 1 and 2, as well as matching of clusters 3 and 4, was verified by using correlation as illustrated in Figure 2. Clusters within state are different because the pattern of their derivatives has opposite signs. The DMN plays an important role in the clusters of State A, with DMN DASWC having an opposite sign compared to other DASWCs. The SBC DASWC can highlight a different sign compared to the other DASWCs through the brain for State B.

**Figure 2.**
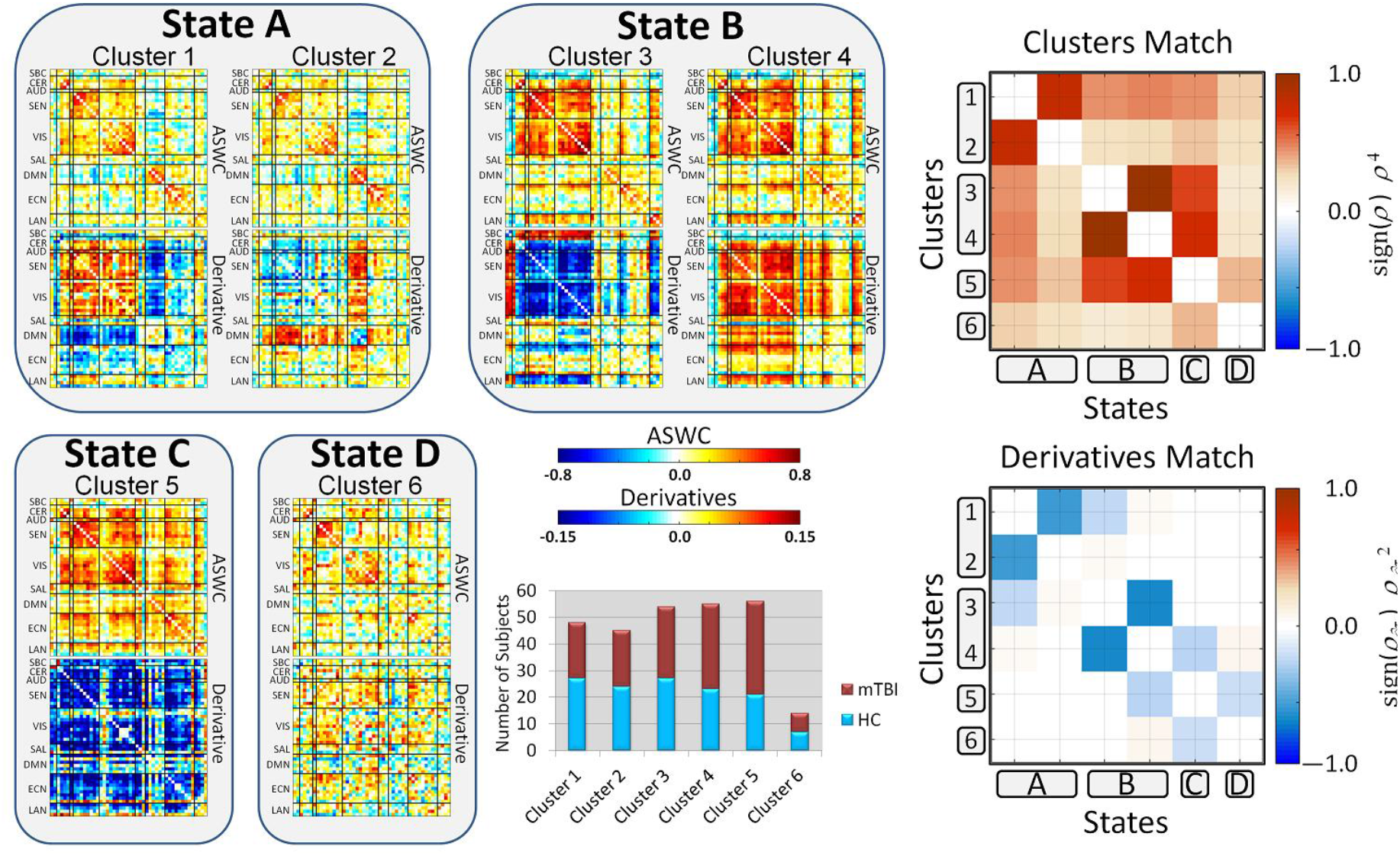
Connectivity patterns of ASWC and their derivatives identified as six different clusters using k-means clustering. The top matrix of each centroid displays correlation patterns (ASWC) and the bottom one their derivatives. As previously defined in (Vergara *et al*., 2018), brain regions of interest were grouped in the functional domains: subcortical (SBC), cerebellum (CER), auditory (AUD), sensorimotor (SEN), visual (VIS), salience (SAL), default mode network (DMN), executive control network (ECN) and language (LAN). Visually, there are some noticeable pairings of clusters with similar correlation patterns having derivative patterns of opposite sign. The pairings are (1, 2) and (3, 4). To verify cluster pairings we correlated cluster patterns, represented by *ρ*, and made sure that ASWC patterns were highly correlated, while their derivatives were highly anti-correlated (derivative of opposite sign).

State C and State D are the two states with only one cluster. Their connectivity and derivative patterns are not matched with other states. There is no distinct functional domain to be highlighted in these states. The functional connectivity pattern of State C is a generalized strong connectivity throughout the whole-brain with a generalized strong negative derivative. This suggests a tendency for the brain to diminish the highly connected brain probably looking for a homeostatic equilibrium. The connectivity pattern of State D indicates a generalized weak derivative with strong connectivity within domains and weak connectivity between domains. State D was scarcely observed, which gives us reason to believe State D does not have a strong influence in this sample.

The temporal sequence of brain patterns is an outcome of the k-means clustering assigning a cluster index to each time point. It is then possible to determine the order of appearance of clusters which have been indexed from 1 to 6. For example, a temporal sequence of clusters *X* = [1, 1, 1, 2, 2, …] defines an instance where the previous cluster *X*_*prev*_ = 1 and the next is *X*_*next*_ = 2. Instances where the next cluster is not the same as the previous are temporal transitions from a cluster to the next and can be mathematically written as. {*X*_*next*_|*X*_*next*_ ≠ *X*_*prev*_}. Several subjects showed oscillations in their cluster sequence, for example *X* = [1, 1, 1, 2, 2,, 2, 2, 1, 1, 1, 1, 2, 2, 2 …] which in the hypothetical picture of Figure 1 indicates an attractor. Since clusters 1 and 2 belong to the same state, this behavior keeps dynamic variations attracted to the functional connectivity pattern of clusters 1 and 2, or State A as previously defined. We estimated the probability of every transition P{*X*_*next*_|*X*_*next*_ ≠ *X*_*prev*_}. through their frequency of occurrence within a given sample group. Figure 3 displays the transition-matrices-results along with the significant differences.

**Figure 3.**
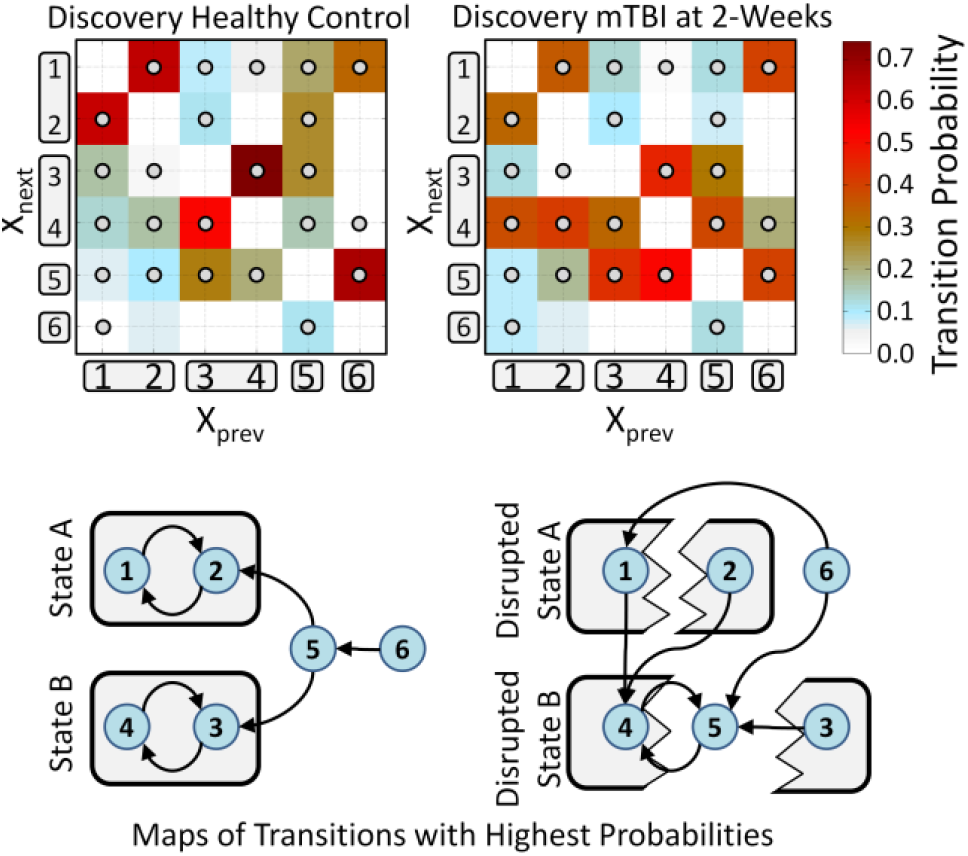
Cluster ***X*** transition probability matrices **P**{***X***_***next***_|***X***_***next***_ ≠ ***X***_***prev***_} and most probable transition structures. Clusters have been grouped using the matching from Figure 2. Significant differences between transition probabilities of HC and mTBI are indicated by a small circle on each colored matrix element box. This figure focuses on State A and State B displaying an attractor behavior. The diagrams for HC show a structure in which State A and State B are attractors since the included clusters transit to each other with highest probability. This attractor-behavior gets disrupted in mTBI patients for State A and State B.

### 2.2. Four Months Follow Up

A total of 23 mTBI scans of subjects from the discovery cohort were taken four months post-injury (mean 111.3 ± 14.2 days post-injury). These scans were not part of the initial dFNC analysis with 96 scans and they were separately analyzed. We extracted temporal information from the available ROIs using a group-information-guided independent component analysis (GIG-ICA) (Du and Fan, 2013). Similar to the previous analysis, ASWC and DASWC estimations were concatenated at each time point. Data at each time point was correlated with each of the six available centroids to determine the nearest cluster. Notice that this procedure does not require k-means since centroids are already known from the previous k-means analysis.

### 2.3. Replication Cohort

The replication cohort consisted of 20 HC (mean age 36 (18-61), 70% male), 54 mTBI patients (mean age 37 (19-64), 67% male) scanned on average at 4 weeks post-injury, and 49 mTBI patients also scanned 3 months post trauma (mean age 37 (19-62), 67% male) (van der Horn *et al*., 2017; van der Horn *et al*., 2018). We applied the same NOIs from the discovery sample. Temporal information of these NOIs was obtained using the GIG-ICA method (Du and Fan, 2013) applied independently to each subject in the replication cohort. This procedure allowed applying ASWC and DASWC to estimate time dependent matrices of dynamic functional connectivity and their derivatives.

### 2.4. Statistical Analysis

The group test performed in this work focuses on transition probabilities which consisted on a single assessment per group. To overcome this problem, we employed a bootstrap procedure using 10k iterations. A random resample with replacement was performed for each subject group (one for controls and one for mTBI groups) and for each iteration. A group difference is then calculated for the current iteration after estimating transition probabilities from the resample groups. Finally, we estimated the probability that group difference is larger than zero using a frequency estimate from the 10k bootstrap iterations.

## 3. Results

### 3.1. Discovery Cohort

The most notable pattern is that opposite derivative cluster pairs (1, 2) and (3, 4) transitions to each other with the highest probability in healthy controls. This type of transition corresponds to the same dynamic state which is indicated by boxes grouping cluster groups (1, 2) and (3, 4) in Figure 3. State A and State B can be identified as two attractor states in the HC group. The observation for mTBI patients is different because clusters 1 and 2 go to 4 with the highest probability, thus indicating a disruption in the dynamic behavior of brain State A. In addition, clusters 3 and 4 go to cluster 5 with the highest probability characterizing a disruption of the dynamic attractor State B. The mTBI patients thus suffer from disruption in both attractor states identified for healthy brains.

### 3.2. Four Months Follow Up

Transition probabilities are displayed in Figure 4. Cluster transitions present the same orbits observed in healthy controls. At four months post-injury, the disrupted attractor behaviors of State A and State B that were found at 2 weeks, are restored (Figure 4). These results suggest timeline of initial injury related disruption of brain state attractors with subsequent recovery of attractor behavior.

**Figure 4.**
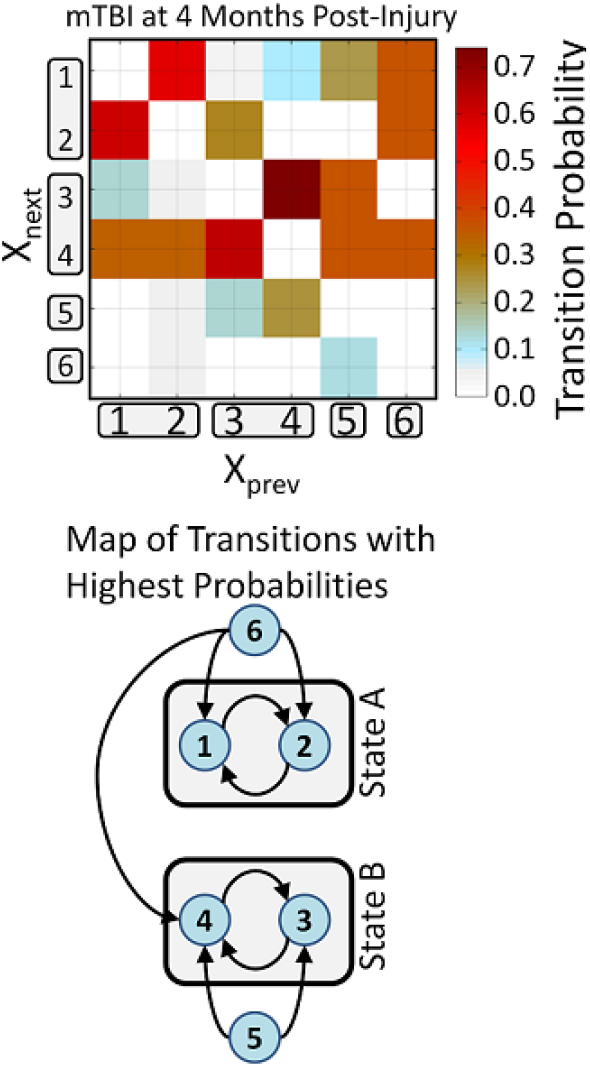
Cluster transitions at 4 months post-injury. The orbits of the two attractor states are intact. This mTBI transition map shows similar State A and State B transitions to those found in the healthy control map of Figure 3. Attractor states of mTBI patients have been restored at four months when compared to the mTBI transitions at 2 weeks.

### 3.3. Replication Cohort

The set of centroids obtained from the discovery data set in Figure 2 was used in the replication data set. At each time point, a matching procedure between centroid and dynamic connectivity matrices was performed by selecting the nearest centroid. Specifically, the centroid exhibiting higher correlation with the matrix at a specific time-point was assigned to that time point. The rest of the analysis is the same as in the discovery data set. As depicted in Figure 5, the HC samples in this cohort replicated the dynamic characteristics of State A and State B as they are again behaving as attractors. Two hypotheses were verified here: first the existence of attractors, and second, that attractor-like behavior is a characteristic of a healthy brain. Next, we can observe that mTBI samples at 4 weeks post injury exhibit a disruption in attractor State B consistent with the hypothesis that brain injury might change the behavior of attractors. Finally, all attractors at 3 months post injury behave similar to HC transition dynamics. This result may suggest that healthy brain connectivity dynamics is restored after three months post injury.

**Figure 5.**
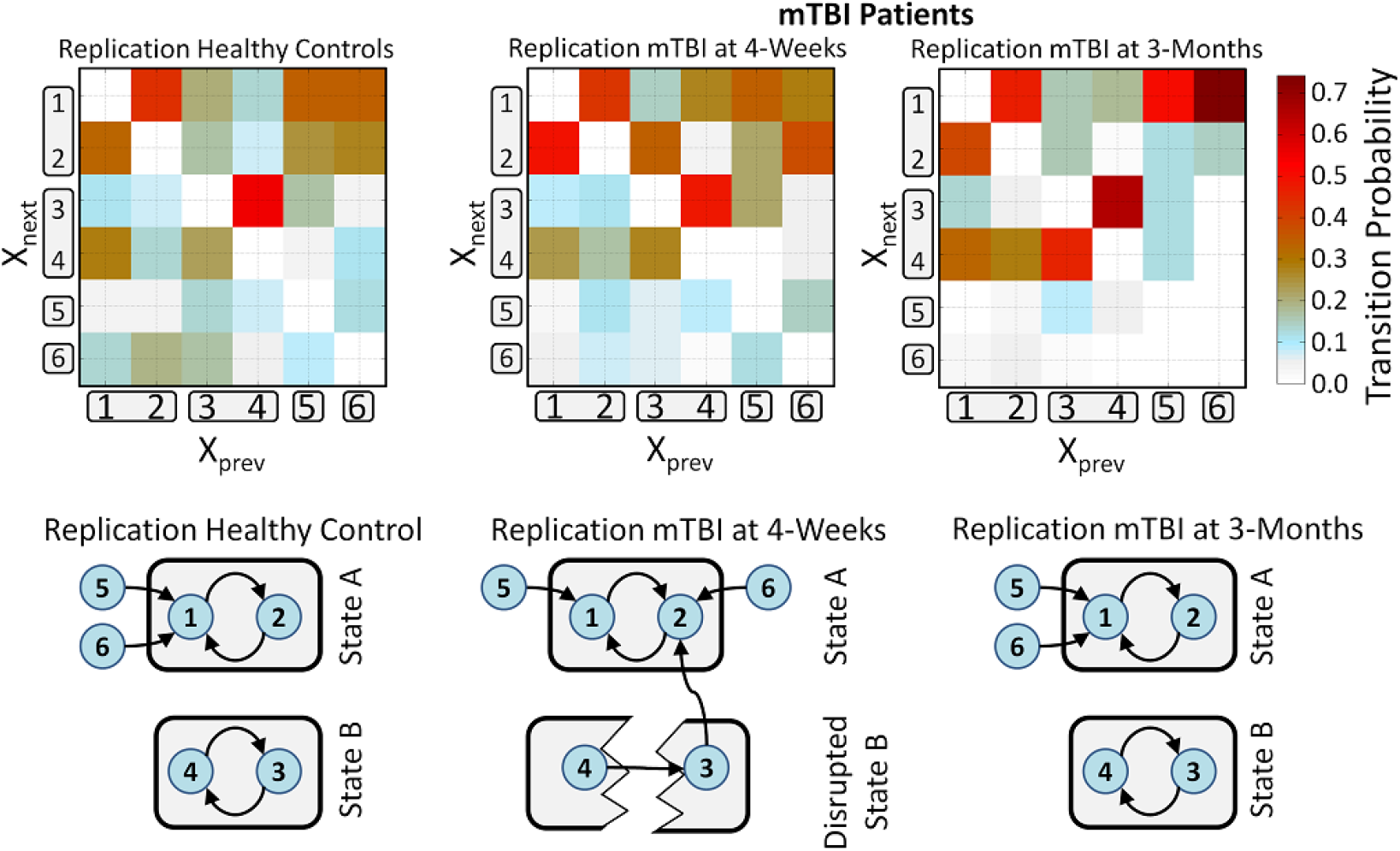
Replication of cluster ***X*** transition probability matrices **P**{***X***_***next***_|***X***_***next***_ ≠ ***X***_***prev***_} and most probable transition structures. Clusters have been grouped using the matching from Figure 2. In this cohort, resting-state fMRI scans (eyes-closed) were obtained at two time points: Time Point 1 at 4 weeks average and Time Point at 3 months average. This figure focuses on State A and State B displaying an attractor behavior. Attractor behavior of State A and State B (as shown in Figure 3) was replicated in these HC. Regarding mTBI, State B was disrupted at Time Point 1, but the transition scheme at Time Point 2 shows restoration of this dynamic brain-state.

## 4. Discussion

This work demonstrates the existence of attractors as an important characteristic of brain functional dynamics, as well as disruption of these attractors due to brain trauma. Presented evidence indicates that healthy whole brain connectivity varies as a function of time, in an oscillatory manner orbiting an attractor centroid. Whole brain connectivity was based on correlation estimates among NOIs distributed throughout the brain. Thus, whole brain connectivity resides in a space with as many dimensions as the number of available correlations. An attractor exists when multi-dimensional time-varying connectivity exhibits relatively small oscillatory changes without a major shift in the overall connectivity pattern. Average dynamic connectivity is then at the center of the multi-dimensional orbit, and useful for representing the orbital functional connectivity structure. Multi-dimensional orbiting is consistent with the hypothesized quasi-static behavior within a dFNC-state as formerly described by Allen et al (Allen *et al*., 2014). Regarding non-healthy brain dynamics, we verified in two different cohorts that brain injury disrupts healthy attractor behaviors. The probability of dynamic connectivity completing a healthy orbit is small when a particular attractor is disrupted. Two striking observations were made in the mTBI discovery cohort: 1) there is a reduced orbiting behavior about centroids (attractor disruptions); and 2) there are attractors present which are not observed in healthy controls (abnormal attractors). Attractor disruption was also observed in the replication mTBI group at four weeks post injury. This disruption seems to have been resolved with time as healthy, 3 and 4 months post injury data exhibited the same attractor characteristics.

The full set of results obtained in this work and presented in Figure 3, Figure 4 and Figure 5 has several implications that may be difficult to notice through the transition matrices. One important result is that orbiting transitions within State A (between clusters 1 and 2) and within State B (clusters 3 and 4) were highly similar for the two HC groups. Once brain connectivity gets configured in cluster 1, results demonstrate that it will vary with time until reaching the configuration of cluster 2 with the highest probability, after which it will return to cluster 1 with the highest probability. Connectivity dynamics will orbit within State A (oscillating between clusters 1 and 2) until a yet not determined cause moves connectivity away from the orbit of State A. One might wonder where dynamic connectivity moves after abandoning attractor State A. Results from Figure 3 and Figure 5 indicate that it will follow a path of clusters and state transitions until it falls into another state orbit. That is, it will return to orbit State A or will be attracted towards orbiting State B comprising clusters 3 and 4. Non-attractor states C and D are part of transient pathways in-between the main-attractors. Attractor brain dynamics in this work can thus be modeled as follows: Functional connectivity is not static during a resting fMRI experiment; instead it moves through several quasi-static functional connectivity states. Quasi-stationarity means there is a specific average connectivity pattern around which there are specific patterns of temporal connectivity variations. One of the observed time-varying patterns describes an orbit around a center described by averaging all points in the orbit. The other type of time-varying patterns does not describe orbits occurring in between orbits. This behavior is depicted in Figure 6 for three different orbits selected from real data and their conceptual summary. For further clarity, a supplementary movie displaying time varying connectivity from real data is provided with this publication. Analyzed data showed evidence to consider this orbiting behavior as healthy brain dynamics.

**Figure 6.**
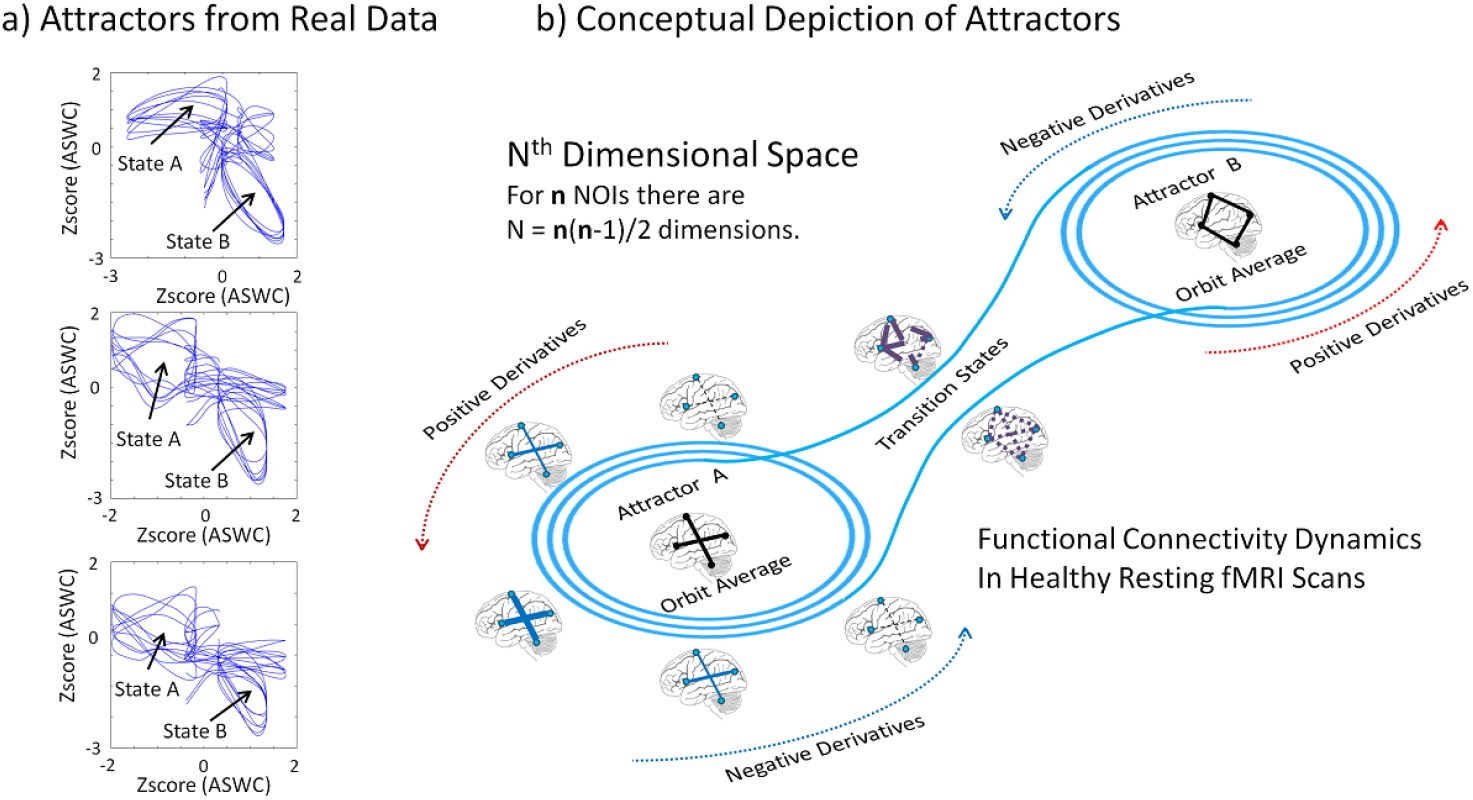
Attractors in dynamic brain connectivity. a) Selected data of a 10 minute scan from a healthy subject moving from the two orbits State A and State B. Dynamic connectivity was assessed using an average sliding window correlation and Z-scored before plotting. These plots show some few examples selected from the vast amount of available data. b) Conceptual depiction of healthy functional connectivity dynamics based on the real data attractor in a). Dynamic functional connectivity varies in an N^th^ dimensional space where each dimension corresponds to an available correlation. For a total of n brain NOIs there are N=n(n-1)/2 different correlations corresponding to all possible NOI pairs. The figure illustrates two orbits where the connectivity increases (positive time derivatives) and decreases (negative time derivatives) in an oscillatory manner. Jumping from one orbit to another may temporally elicit transitional connectivity states. This pattern was observed in healthy controls and described by the state transitions with highest probability in discovery and replication datasets.

The resting state brain-dynamics-model described in the previous paragraph is consistent with findings and hypotheses described in the literature. Functional connectivity in State A follows an expected pattern shown in resting state experiments, where the DMN is negatively correlated with other brain regions, but strongly internally connected (Greicius *et al*., 2003; Buckner *et al*., 2008; Raichle, 2015). This pattern has been linked to self-referential and mindfulness processing due to the strong influence of ventro-medial prefrontal cortex and the posterior cingulate (Raichle, 2015). In addition, fMRI experiments during a mindful breathing task has shown that subjects with higher mindfulness disposition prefer connectivity patterns similar to State A, consisting of anti-correlations between the DMN and networks that are important for focused attention, with the latter being highly inter-connected (Mooneyham *et al*., 2017). Thus, a disruption of State A in patients with mTBI may lead to a reduced ability to be mindful and to engage in attentive mental processes. It is likely that neuroplasticity attempts to reinstate this broken attractor, and this type of brain healing in mTBI may even be stimulated by mindfulness training rehabilitation techniques (Evans, 2012; Johansson *et al*., 2012; Niraj *et al*., 2018).

The second main attractor-state, State B, exhibits stronger connectivity within sensorial and motor domains VIS, AUD and SEN, all of which are negatively correlated with the SBC domain composed of basal ganglia and thalamus areas. Our results show that this state also gets disrupted after mTBI, with non-attractor state C (cluster 5: which contains positive functional connections between subcortical regions and cortical domains) becoming an attractor with cluster 4 (which contains negative correlations between subcortical and cortical regions), forming a new dynamic brain state. Patients with mTBI often report increased sensitivity to light, sound, pain, and patients are easily overwhelmed by cognitively challenging stimuli that are encountered during everyday life (de Koning *et al*., 2016). These symptoms might be reflected by the change in state B, possibly indicating that processing of various exteroceptive stimuli is increased. According to the study by Mooneyham et al. (Mooneyham *et al*., 2017) there is a second possible mindset in addition to the mindful/attentive mindset, namely one representing mind wandering which is associated with a state of high connectivity between DMN and networks for focused attention. Another study has shown that higher connectivity between these networks is associated with lower mindfulness ability (Doll *et al*., 2015). As cluster 5 in our current study also shows increased functional connections of the DMN and ECN with other brain regions, such as the VIS, AUD and SEN domain, it could be argued that the change in state B encompasses increased mind-wandering, and perhaps even rumination, in response to increased bottom-up processing of exteroceptive stimuli. These theories are supported by a study by Sours et al., that revealed increased functional connectivity between sub-thalamic regions and various cortical areas including those of sensory processing and the default mode network in patients with mTBI (Sours *et al*., 2015). Other studies have also shown changes in connectivity strength in thalamocortical areas, and increases in connectivity in a network of subcortical structures including thalamus, hippocampus and amygdala after mTBI (Tang *et al*., 2011) (Iraji *et al*., 2015). As cortico-striatal-thalamic-cortical loops are involved in mental processes such as cognition, learning and memory, alterations of attractor dynamics in state B may also have detrimental effects on cognitive processes, explaining some of the known impairments and symptoms in mTBI, such as mental fatigue (Helie *et al*., 2015; Makino *et al*., 2016; Shine *et al*., 2016) (Dikmen *et al*., 2009). The third mindset described by Mooneyham and colleagues represents a transitional mindset. The hypothetical existence of transitional mindsets is well supported by our hypotheses and our results, since non-attractor states (State C and State D) seem to function as temporal bridges between attractor states, and might even form new brain states after neural perturbations such as mTBI.

It has to be realized that there is a possibility that deviations from healthy attractor states after mTBI in fact already reflect, or partly reflect, recovery processes that are needed to return to a healthy brain state. It has been theorized that changes in functional connectivity after traumatic brain injury, especially during the subacute stage, are the natural brain reaction to the traumatic insult (Tang *et al*., 2011; Vergara *et al*., 2018). A regeneration mechanism gets triggered by axonal damage of white mater and thalamic projection fibers (Little *et al*., 2010). Neuroplasticity tries to compensate by regenerating injured axons, through branching of unaffected axons or remodeling the neural circuitry (Navarro, 2009; Chipman, 2016). Traumatic brain injury causes alterations in hemodynamics characterized by hypoperfusion mainly in the thalamus, but also in cortical regions (Kim *et al*., 2010). These healing mechanisms likely change the dynamic interaction of functional connectivity. The current study shows that both the attractors State A and State B are disrupted at approximately 2 weeks post injury. This may suggest that both mindfulness and wandering mental processes, as described by Mooneyham et al., are impaired as a result of the different neuroplastic events necessary to restore normal brain function (Mooneyham *et al*., 2017). Furthermore, an abnormal attractor between clusters 4 and 5 appears to replace State B (see Figure 3), suggesting that brain dynamics try to retain an attractor like-behavior instead of a full chaotic disruption. Results at 4 weeks (late sub-acute) and 3 months (early chronic) describe the evolution of brain healing after mild TBI. Attractor State A was restored at 4 weeks while attractor State B was subsequently restored at 3 months.

At this point, it is possible to draw a simplified timeline (by disregarding state C and D) of dynamic disruption and restoration after mTBI as illustrated in Figure 7. Immediately after injury, brain dynamics enters partial chaotic behavior where abnormal attractor dynamics occurs. The brain eventually tries recovering normal attractor dynamics, which is completed at three months post-injury. This timeline successfully describes results from discovery and replication experiments, despite being performed in different cohorts.

**Figure 7.**
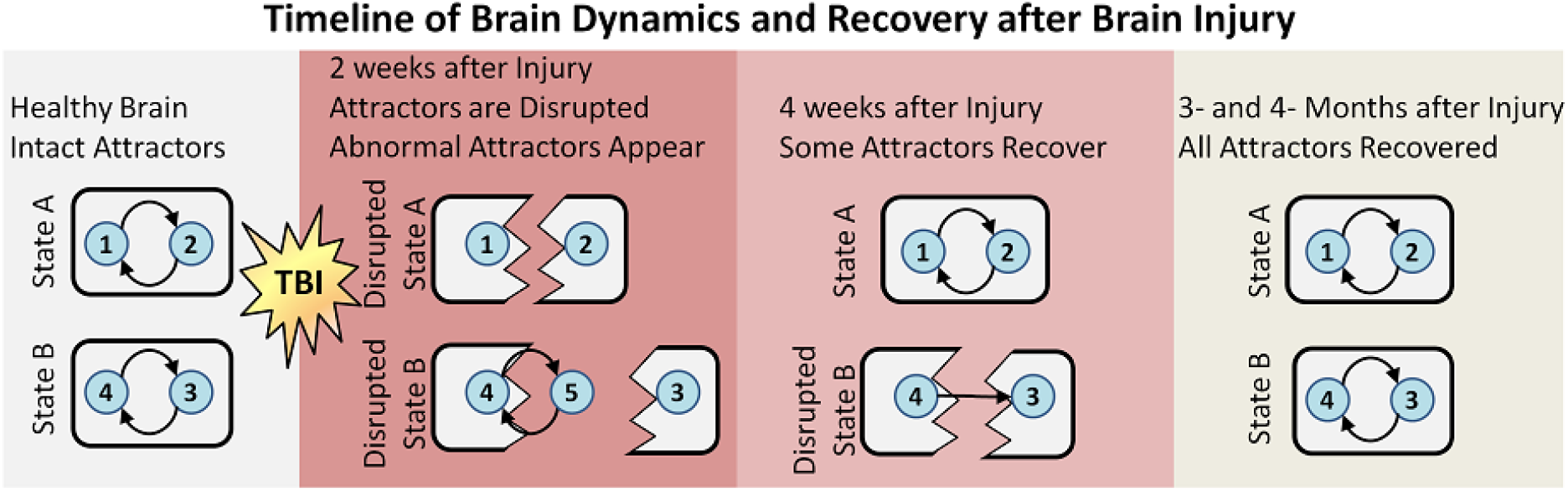
Devised timeline of brain dynamic behavior after brain injury. This figure is based on results from Figure 3 and Figure 5. Healthy brains have a dynamics functional connectivity that orbits around specific connectivity patterns configured as attractors. Brain injury disrupts the orbits and functional connectivity dynamics enters an abnormal phase. Some attractors recover after four weeks. Complete attractor behavior was observed after three and four months.

Discovery results presented here are from mTBI subjects in the subacute stage. The observations made are then consistent with the establishment of temporary neurodegenerative mechanisms short after brain injury. The replication result allowed us to verify the healthy condition as well as observing longitudinal dysfunctions after mTBI. Although attractor states behavior was replicated, results for the non-attractor states were slightly different which could be partly explained by the heterogeneity of mTBI cases. Our experimental design could not fully replicate or explain details of these non-attractor states and more research is necessary to elucidate their relevance in mTBI. As is true for any group analysis, it is still difficult to particularize the results for specific subjects. Results can only explain effects in a general sense. Part of this problem relies in the fact that not all clusters were observed in all subjects. While using bootstrapping helped in overcoming this limitation, a future study design could minimize this problem, possibly by taking several fMRI scans on different days for the same subject at the same stage of mTBI. This will increase the possibility of detecting the cluster transitions defined in this work.

## Data Availability

Data has not been yet available for public release.

## 5. Funding

This work was funded by the following NIH grants: R21NS064464/R21NS064464/R01NS098494 to A.M. and P20GM103472/1R01EB006841/R01REB020407 and by the National Science Foundation (#1539067) to V.C. For the replication cohort, funding was obtained from the Dutch Brain Foundation (grant number: Ps2012-06; to J.N.). No competing financial interests exist.

## Notes

### Competing Interest Statement

The authors have declared no competing interest.

### Author Declarations

All relevant ethical guidelines have been followed and any necessary IRB and/or ethics committee approvals have been obtained.

Any clinical trials involved have been registered with an ICMJE-approved registry such as ClinicalTrials.gov and the trial ID is included in the manuscript.

